# GPT-4 outperforms ChatGPT in answering non-English questions related to cirrhosis

**DOI:** 10.1101/2023.05.04.23289482

**Authors:** Yee Hui Yeo, Jamil S. Samaan, Wee Han Ng, Xiaoyan Ma, Peng-Sheng Ting, Min-Sun Kwak, Arturo Panduro, Blanca Lizaola-Mayo, Hirsh Trivedi, Aarshi Vipani, Walid Ayoub, Ju Dong Yang, Omer Liran, Brennan Spiegel, Alexander Kuo

## Abstract

**Background and Objectives:** Artificial intelligence is increasingly being employed in healthcare, raising concerns about the exacerbation of disparities. This study evaluates ChatGPT and GPT-4’s ability to comprehend and respond to cirrhosis-related questions in English, Korean, Mandarin, and Spanish, addressing language barriers that may impact patient care.

**Methods:** A set of 36 cirrhosis-related questions were translated into Korean, Mandarin, and Spanish and prompted to both ChatGPT and GPT-4 models. Non-English responses were graded by native-speaking hepatologists on accuracy and similarity to English responses. Chi-square tests were used to compare the proportions of grading between ChatGPT and GPT-4.

**Results:** GPT-4 showed a marked improvement in the proportion of comprehensive and correct answers compared to ChatGPT across all four languages (p<0.05). GPT-4 demonstrated enhanced accuracy and avoided erroneous responses evident in ChatGPT’s output. Significant improvement was observed in Mandarin and Korean subgroups, with a smaller quality gap between English and non-English responses in GPT-4 compared to ChatGPT.

**Conclusions:** GPT-4 exhibited significantly higher accuracy in English and non-English cirrhosis-related questions, highlighting its potential for more accurate and reliable language model applications in diverse linguistic contexts. These advancements have important implications for patients with language discordance, contributing to equalizing health literacy on a global scale.

## Introduction

The use of Artificial intelligence (AI) is growing in the field of medicine with the advent of new and innovative tools that can refine various aspects of patient care, from diagnosis to long-term management. However, it is important to identify and prevent the exacerbation of systemic racial, ethnic, and sex disparities in healthcare with the utilization of AI tools in medicine.[1] ChatGPT is an innovative natural language processing tool that can comprehend complex inquiries and provides easy-to-understand conversational responses that are seemingly knowledgeable.[2] ChatGPT and GPT-4 were released in November 2022 and March 2023. Despite its recent release, there is a rapidly growing body of evidence showing its remarkable ability to answer clinically related questions.[3, 4, 5]

Cirrhosis is a complex condition that constitutes 2.4% of worldwide deaths and requires meticulous care to prevent complications.[6, 7, 8] Language barriers could impact the quality of care, as patients with restricted English proficiency could experience barriers to receiving adequate medical attention. Patients who had language discordance with their healthcare provider experienced an increased likelihood of patient dissatisfaction and worse outcomes.[9]

ChatGPT 4 has shown a significant improvement in cross-lingual comprehension and translation.[10] This has the potential to diminish the language barrier and address the racial disparity. In this study, we examined the models’ ability to understand and reply to cirrhosis-related questions in Korean, Mandarin, and Spanish while comparing its performance to English. We also compared the accuracy of responses between ChatGPT and GPT-4.

## Method

Frequently asked patient questions about basic knowledge of cirrhosis were used from our previous article to ensure consistent evaluation of ChatGPT’s capability.[3] These questions were obtained from reputable healthcare organizations and patient support groups. A total of 36 questions in the domain of basic knowledge were included. Each question was translated into Korean, Mandarin, and Spanish. Questions in different languages were independently prompted to both ChatGPT and GPT-4 models to obtain a response.

### ChatGPT and GPT-4

ChatGPT is a natural language processing (NLP) model that has been designed as a variant of GPT-3.5 LLM (Large Language Model). It is trained on a vast dataset of information collected from various online sources up to September 2021, such as books, websites, and articles. ChatGPT is trained using Reinforcement Learning from Human Feedback (RLHF), which incorporates human feedback and correction to generate coherent and contextually appropriate responses [11]. This allows for responses to be concise, understandable, and well-formulated. Users can input any prompt into ChatGPT, which uses the information stored in its database to generate a response.

GPT-4 is the successor of ChatGPT. Although exact technical specifications of GPT-4 have not been publicly disclosed, GPT-4 boasts superior performance over its predecessor, and displays advance reasoning capabilities. In 24 of 26 language versions tested, GPT-4 has far outscored ChatGPT in the Massive Multitask Language Understanding (MMLU) examination, which covers 57 different subjects [12]. GPT-4 is also able to better detect inappropriate prompts and regulate its answers. Being a multimodal LLM, it can receive both image and text prompts, whereas ChatGPT can only receive text prompts.

### Grading

Non-English responses were collected and subjected to two methods of grading: 1) the accuracy of each response, 2) similarity to the English response.

These responses were graded by hepatologists who are native speakers. The accuracy of each response was graded using the scale: 1) comprehensive, 2) correct but inadequate, 3) some were correct, while some were incorrect, and 4) completely incorrect. Grading was based on the American Association for the Study of Liver Diseases (AASLD) guidelines. Given that the reviewers for each language are different, we did not compare the proportions of the grading across the language directly.

We, instead, performed comparisons between English and non-English responses by having the native speaker hepatologists (all were also proficient in English) assess and compare the accuracy. The comparison was performed using the scale 1) the English response has more accurate explanations than the non-English response, 2) the same level of accuracy, and 3) the English response has less accurate explanations than the non-English response.

### Statistical Analysis

The proportion of responses in each grade for both grading methods was calculated. Chi-square test was applied to compare the accuracy of proportions of grading between ChatGPT and GPT-4. A p-value of >0.05 was considered statistically significant.

Analysis was done with IBM SPSS Statistics (Version 22).

## Results

The workflow of response data collection and assessment is summarized in **Figure 1**. We saw a marked improvement in the proportion of comprehensive and correct answers by GPT-4 compared to ChatGPT across all four languages (p<0.05 for each language) **(Table 1)**. Notably, the responses generated by GPT-4 demonstrated enhanced accuracy and avoided erroneous responses that were evident in ChatGPT’s output. For example, hepatic encephalopathy and hepatic steatosis were used interchangeably in Korean using ChatGPT, which rendered the Korean responses to be completely incorrect in several questions. The Spanish version provided an incorrect definition of heavy drinking, and the Mandarin version suggested the use of anti-coagulation medications to prevent variceal bleeding. However, these mistakes were not found in GPT-4’s responses.

**Figure 1.**
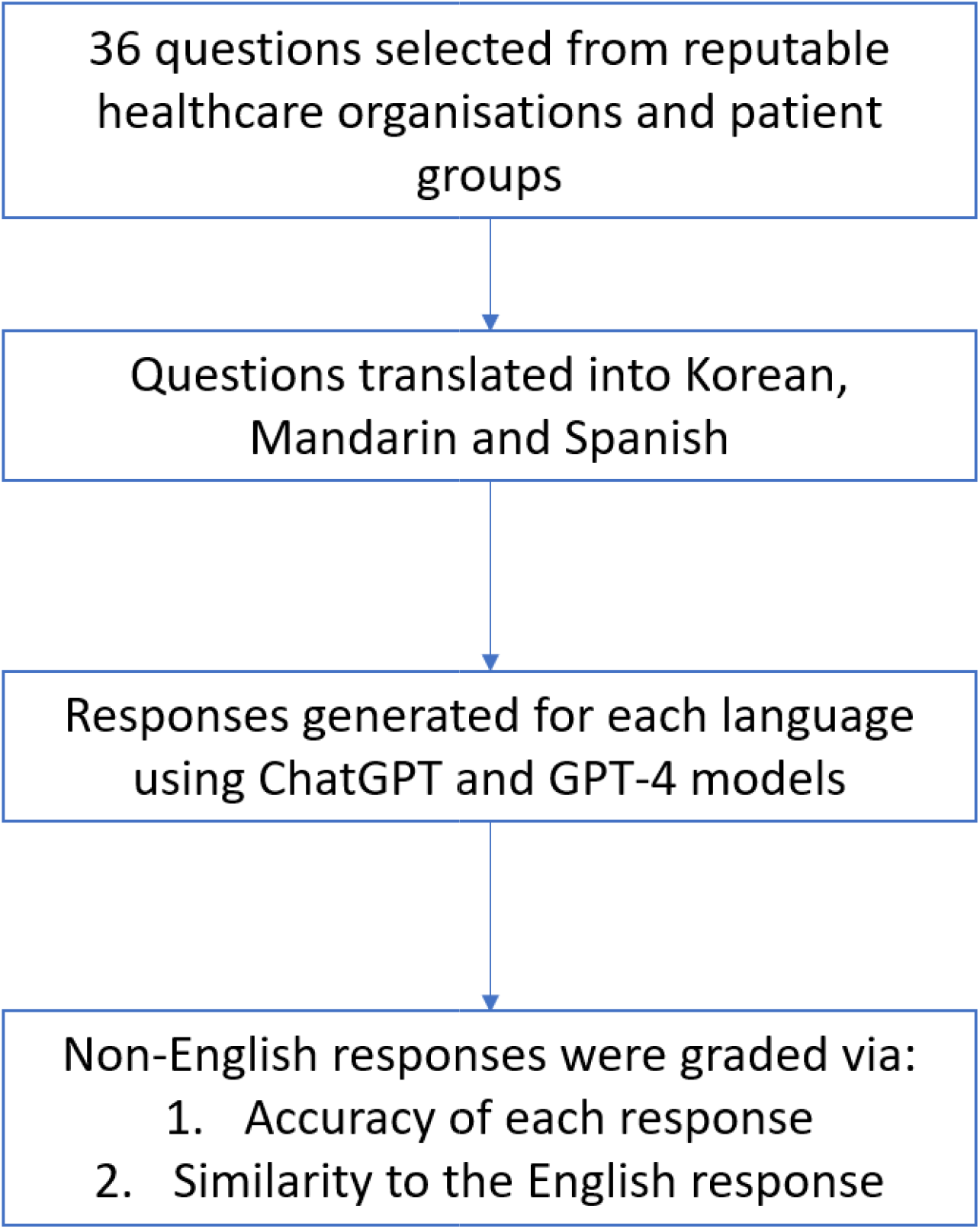
Flowchart of data collection and analysis.

**Table 1.** Grade of responses by GPT-3.5 and GPT-4 to questions related to cirrhosis in different languages.

|  |  | **Comprehensive** | **Correct but inadequate** | **Mixed with correct and incorrect/outdated data** | **Completely incorrect** | **P value\*** |
| --- | --- | --- | --- | --- | --- | --- |
| English | GPT-3.5 | 47.2% | 30.6% | 22.2% | 0.0% | 0.015 |
|  | GPT-4 | 75.0% | 11.1% | 13.9% | 0.0% |  |
| Spanish | GPT-3.5 | 30.6% | 44.4% | 22.2% | 2.8% | <0.001 |
|  | GPT-4 | 69.4% | 11.1% | 19.4% | 0.0% |  |
| Mandarin | GPT-3.5 | 16.7% | 63.9% | 19.4% | 0.0% | 0.01 |
|  | GPT-4 | 38.9% | 55.6% | 5.6% | 0.0% |  |
| Korean | GPT-3.5 | 5.6% | 19.4% | 33.3% | 41.7% | <0.001 |
|  | GPT-4 | 69.4% | 5.6% | 19.4% | 5.6% |  |

\*p-value of the comparison in accuracy between the responses of GPT-3.5 and GPT-4

Furthermore, while the English responses consistently provided more comprehensive explanations than their non-English counterparts for both ChatGPT and GPT-4, significant improvement was observed in both Mandarin (0.018) and Korean (<0.001) subgroups (**Table 2**). For responses generated by ChatGPT, although many responses were factually correct, they were inadequate and very succinct, with less explanation, resulting in less abundant information compared to the English version. Compared to the responses generated by ChatGPT, a larger proportion of GPT-4’s responses in Mandarin and Korean were considered to possess comparable levels of accuracy.

**Table 2.** Difference in the accuracy of GPT-3.5 and GPT-4 in response to English vs. non-English cirrhosis-related questions.

|  |  | **Having less accurate explanations than the English version** | **Similar level of responses** | **Having more accurate explanations than the English version** | **P value\*** |
| --- | --- | --- | --- | --- | --- |
| Spanish | GPT-3.5 | 41.7% | 44.4% | 13.9% | 0.096 |
|  | GPT-4 | 41.7% | 55.6% | 2.7% |  |
| Mandarin | GPT-3.5 | 63.9% | 30.6% | 5.6% | 0.018 |
|  | GPT-4 | 36.1% | 52.8% | 11.1% |  |
| Korean | GPT-3.5 | 91.7% | 8.3% | 0.0% | <0.001 |
|  | GPT-4 | 27.8% | 72.2% | 0.0% |  |

\*p-value of the comparison between GPT-3.5 and GPT-4 in the accuracy of English vs. non-English languages

## Discussion

In this study, GPT-4 demonstrated a significantly higher accuracy in both English and non-English questions related to cirrhosis. We also showed a smaller quality gap between the English and non-English responses in GPT-4 compared to ChatGPT, as there was a significant increase in the proportions of responses with a similar level of accuracy between English and Mandarin, and Korean.

The negative impact of language barriers on healthcare outcomes has been previously described. A meta-analysis of 14 studies which included 300,918 participants, revealed that language barriers in healthcare might lead to miscommunication between healthcare providers and patients, leading to reduced satisfaction for both parties, as well as a decline in the quality of healthcare delivery and patient safety.[9] These disparities have also been demonstrated among patients admitted to the hospital where patients with limited English proficiency experienced higher rates of adverse events compared to patients proficient in English.[13] A qualitative study of immigrants with limited proficiency in English highlighted concerns among patients, specifically among those with chronic diseases which require the availability of after-visit resources to aid in education and compliance with medical directions.[14] Furthermore, studies examining online Spanish medical information found sources to either be non-readable with higher than recommended reading levels or of low quality.[15, 16, 17] While we do not advocate for the use of natural language models in patient education outside of the care of a licensed healthcare professional, this rapidly evolving technology may serve as an easy-to-access and highly comprehensible adjunct information source for patients.

Understanding the mechanism by which language learning models can comprehend and respond in multiple languages is important in assessing their performance but also their limitations. Cross-lingual machine reading comprehension can be achieved through multilingual pre-training, which is a technique utilized by language models such as Bidirectional Encoder Representations from Transformers (BERT) and Generative Pre-trained Transformer (GPT).[18, 19] These methods allow the language model to understand words and phrases in different languages by learning in a unified semantic space, facilitating the transfer of knowledge across languages.

GPT-4 outperformed ChatGPT in responding to non-English questions related to cirrhosis. This could be explained by the fact that GPT-4 was trained using a larger pre-trained dataset with more examples of text in various languages.[20] Regarding pre-processing of data, GPT-4 has improved tokenization and handling of unique characters found in different languages. Moreover, GPT-4 is equipped with enhanced training techniques, including transfer learning, zero-shot or few-shot learning (as explained above), multilingual data sampling, and cross-lingual pre-training, allowing it to comprehend and answer non-English questions more effective than ChatGPT.

In conclusion, we demonstrated a significant improvement in GPT-4’s ability to comprehend and accurately respond to both English and non-English cirrhosis-related questions. The advancements of GPT-4 over its predecessor highlight the potential for more accurate and reliable language model applications in diverse linguistic contexts. These advancements have important implications for patients who have language discordance with their healthcare providers and will contribute to equalizing health literacy on a global scale by delivering accurate and comprehensive explanations.

## Data Availability

All data produced are available online using ChatGPT

## Acknowledgments

GPT-4 was used to paraphrase the part of the manuscript.

